# Hypertension mediated the effects of pericardial adipose tissue on cardiovascular diseases

**DOI:** 10.1101/2024.01.02.24300740

**Authors:** Feng Xie, Peng Cao, Hai-bo Tang, Zhi Luo, Shai-hong Zhu, Li-yong Zhu

**Affiliations:** Department of General Surgery, The Third Xiangya Hospital, Central South University, No.138, Tongzipo Road, Yuelu District, Hunan 410013 Changsha, China; State Key Laboratory of High Performance Complex Manufacturing, College of Mechanical and Electrical Engineering, Central South University, 932 South Lushan Street, Hunan 410083 Changsha, China

**Keywords:** pericardial adipose tissue, cardiovascular diseases, Mendelian randomization

## Abstract

**Background:** Although previous studies have presented a relevance between pericardial adipose tissue (PAT) and cardiovascular diseases (CVDs), the precise role of PAT in CVDs remains uncertain.

**Methods:** In the Mendelian randomization (MR) research, we extracted instrumental variants significantly correlated with PAT to assess its effects on CVDs. Inverse-variance weighted model was elected as the leading MR analytical method. F-statistic was utilized to assess the intensity of instrumental variants and avert weak-tool bias. Numerous sensitivity analyses were adopted to confirm the credibility of outcomes. The mediated effect of hypertension between PAT and CVDs was estimated in 2-step MR analysis.

**Results:** The MR research demonstrated genetically determined PAT was remarkably correlated with greater risks of cardiovascular disease (OR 1.15; 95% CI 1.09--1.22, P=1.32*10^-6^), heart failure (OR 2.62; 95% CI 1.28--5.34, P=0.008), coronary heart disease (OR 3.53; 95% CI 1.55--8.06, P=0.003), ischemic heart disease (OR 2.31; 95% CI 1.14--4.67, P=0.020), ischemic stroke (OR 3.18; 95% CI 1.63--6.20, P=0.001), and venous thromboembolism (OR 1.02; 95% CI 1.00--1.04, P=0.015). And the correlation was partially mediated by hypertension. Results were verified by sensitivity analysis.

**Conclusions:** Genetic determined PAT was remarkably correlated with additional risks of diverse CVDs. Identified as a mediator, hypertension had a significant influence on the causal association. The correlation between PAT and CVDs deserves further exploration.

**NOVELTY AND RELEVANCE:** *What Is New?:* This was the first Mendelian randomization research to detect the potential causal relationship of genetically determined PTA on multiple CVDs risks and confirm the mediating effect of hypertension on the causal association.

*What Is Relevant?:* The MR research not only estimated the causal effect of PAT on hypertension but also identified the mediating role of hypertension between PAT and CVDs.

*Clinical/Pathophysiological Implications?:* This study revealed PAT promoted CVDs progression and hypertension played an intermediary role, which indicated that PAT might serve as a potential therapeutic target and antihypertensive drugs could be conjunctly used to provide cardiovascular benefits. especially for people with obesity, because they had more PAT accumulation.

## INTRODUCTION

As a leading cause of death, cardiovascular diseases (CVDs) were accountable for almost 19 million fatalities in 2020.^1,2^ CVDs consist of diverse diseases, such as cerebrovascular disease, coronary artery disease (CAD), and ischemic stroke (IS).^1^ And IS can be primarily categorized into three subtypes, involving cardioembolic stroke (CES), large artery stroke (LAS), and small vessel stroke (SVS).^3^ Lots of etiologic factors are responsible for the development of CVDs.^4^ Diabetes mellitus, hypertension, and hyperlipidemia all contribute to CVDs risk.^5-7^ Furthermore, pericardial adipose tissue is also reported as a suspected risk factor for CVDs.^8^

Pericardial adipose tissue (PAT), as a special constituent of body fat, is a unique fat storage surrounding the outermost layer of pericardium.^9^ PAT is divided into paracardial fat, epicardial fat, and perivascular fat from outside to inside.^10^ Owing to its proximity to heart, PAT has been intuitively assumed to have a vital influence in the pathobiology of CVDs.^11^ The lack of an anatomical barrier allows crosstalk between PAT and the contiguous myocardium. Although one of PAT’s roles is mechanical protection and energetic support,^12,13^ over-expanding PAT turns into a deleterious pro-inflammatory character from a beneficial anti-inflammatory role through paracrine and vasocrine.^9,14^ Amounts of observational studies already demonstrated that PAT is positively correlated with higher risks of hypertension,^15^ heart failure (HF),^16^ atrial fibrillation (AF),^17^ CAD,^18^ and stroke.^19^ However, some research revealed that there was inconsistent relationship between PAT and obstructive CAD, and coronary artery calcification extent had no obvious association with increased PAT volume even after adjusting for age and sex.^20,21^

Of note, most of the previous studies are cross-sectional research, which has a weak strength to explain the causal relationship, so recent research challenged the relationship and emphasized longitudinal studies.^20,22^ Besides, traditional observational research is vulnerable to confounders and reverse causation bias. Therefore, causality in the associations of PAT with CVDs is worth further exploration.

Mendelian randomization (MR) is a nascent methodology for epidemiological studies. MR study adopts instrumental variables (IVs) derived from heritable variants that are dependably correlated with potential risk factors to assess the causal correlation between exposure for phenotype and outcome for disease based on a genome-wide association study (GWAS).^23^ Because of the peculiar strength of IVs, MR research is less vulnerable to reverse causation and confounders compared with conventional observational studies.^24^ Fortunately, a recent GWAS identified multiple heritable variants related to pericardial fat.^25^ By utilizing those single-nucleotide polymorphisms (SNPs) as IVs, we here performed MR research to evaluate the genetical effect of PAT on CVDs, including cardiovascular disease (CVD), HF, cardiac arrhythmias (CA), AF, aortic aneurysm (AA), coronary heart disease (CHD), hypertrophic cardiomyopathy (HCM), ischemic heart disease (IHD), myocardial infarction (MI), IS, CES, SVS, LAS, subarachnoid hemorrhage (SAH) and venous thromboembolism (VTE), and to further inquire the mediating role of hypertension on the causal association.

## METHODS

### Study Design

A summarization of the MR design was presented (Figure 1). The MR research was adopted to assess the causal correlation between PAT and CVDs risks by utilizing heritable variants as IVs. And IVs fulfill the following three principal assumptions: (1) directly related to exposure; (2) separate from confounders; and (3) act on the risk of outcomes only through exposure.^26^

**Figure 1.**
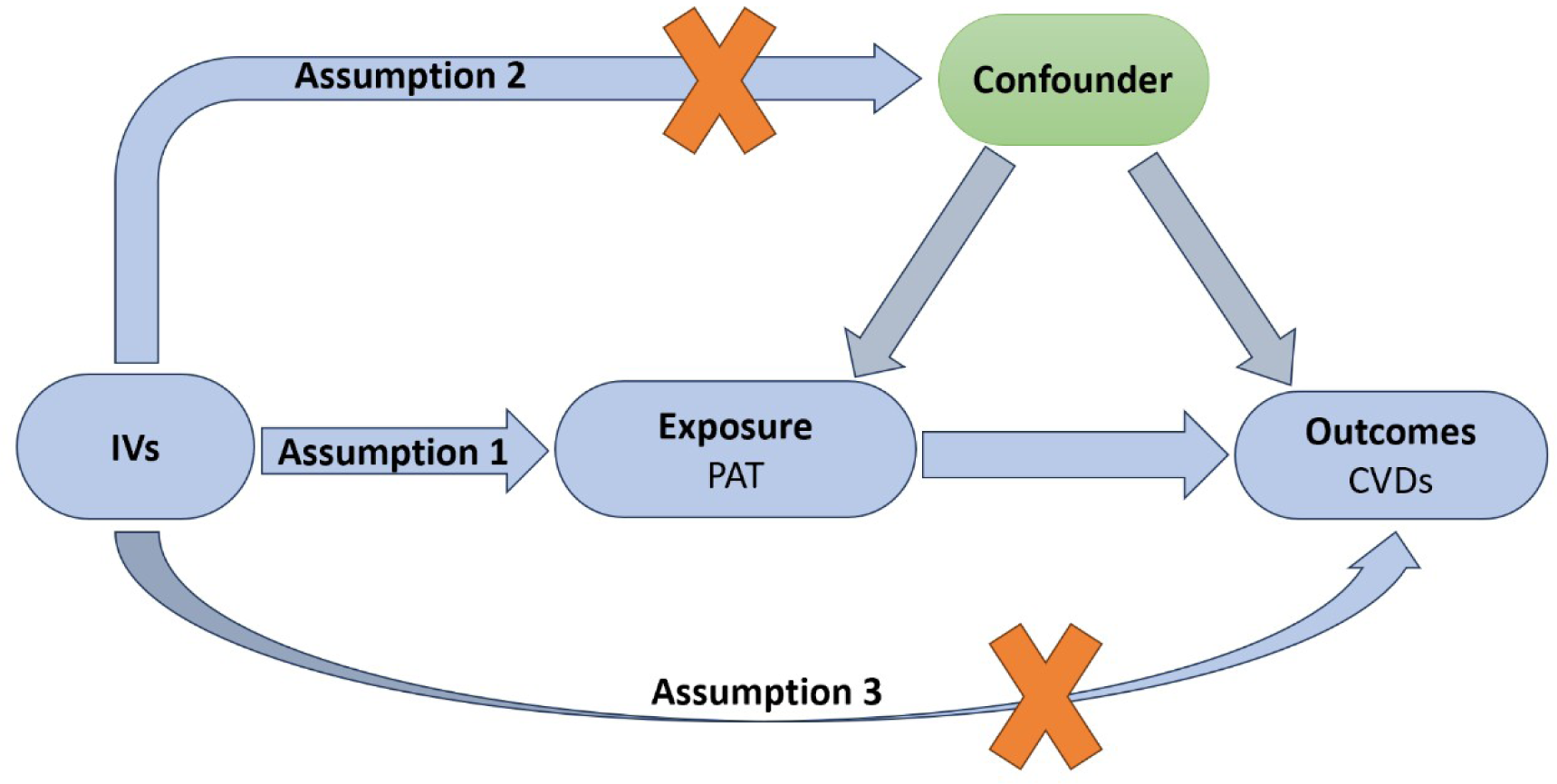
Flowchart of the MR research. Instrumental variants (IVs) fulfill three principal assumptions: (1) directly related to exposure; (2) separate from confounders; and (3) act on outcomes just through exposure. MR, mendelian randomization.

The statistical data for exposure of PAT accumulation were derived from a largest and latest GWAS. PAT was measured by cardiac magnetic resonance images in 28,161 participants with white British heritage.^25^ GWAS datasets for outcomes of all CVDs were retrieved from IEU OpenGWAS project. Statistical data from the MEGASTROKE was acquired for CES, SVS, LAS, and IS.^27^ GWAS datasets for CVD, AF, HTN,^28^ MI, SAH,^29^ and VTE^30^ were derived from UK Biobank. And we acquired most of the GWAS data from Finngen, including CHD, IHD, HCM, AA, CA, and HF.^31^ The details of data sources are described (Table S1). Ethical approval and inform consent of all contributors have been obtained.

### IVs Selection

IVs significantly correlated with PAT were extracted in SNPs from GWAS when the p-value was less than 5×10^-8^. To identify the corresponding linkage disequilibrium (LD) of IVs, we further clumped marked SNPs with 10,000 kb and r^2^ < 0.001 from the 1000 Genomes linkage disequilibrium European samples.^32^ Then we searched each single SNP in PhenoScanner V2 to avoid obvious pleiotropic effects, excluded SNPs corresponding to potential confounding factors and selected the remained SNPs for further analysis.^33^ To estimate the intensity of IVs and control the weak-tool bias, we utilized R^2^ and F-statistic. The calculation method follows: F = (N-2) × R^2^ / (1-R^2^). The R^2^ suggests the percentage of exposure variance construed by IVs, which is computed by the following equation: R^2^=[2×β^2^×EAF×(1−EAF)]/[2×β^2^×EAF×(1−EAF)+2×N×SE^2^×EAF×(1−EAF)].^34^ Here, β, EAF, N, and SE separately refer to the assessed effect, effect allele frequency, sample size, and standard error. Usually, little probability of weak-tool bias is considered if F>10.^34^

### MR Analysis

Inverse-variance weighting (IVW) model was elected as the leading MR analytical method. Although vulnerable to pleiotropy and bias, IVW method can provide the most precise estimated effects.^35^ Thus, in order to infer the association between PAT and CVD, several sensitivity analyses were employed, including MR-RAPS,^36^ MR-Egger,^37^ weighted median,^38^ and MR-PRESSO.^39^ Weighted median analysis gives a permission to estimate causality consistently even though up to 50% of IVs are of invalidity.^38^ MR-Egger was implemented to measure and rectify the possible horizontal multiplicity.^40^ The p-value of intercept greater than 0.05 indicates few remarkable bias of horizontal multiplicity is noted. MR-PRESSO can identify outlying SNPs automatically and exclude outliers to correct MR estimates.^39^ In addition, Q statistics was employed to detect heterogeneity between individual SNP in IVW analysis, which could guide to an appropriate method.^38^ When the p-value for Q statistic is smaller than 0.05, which indicates the possibility of heterogeneity, IVW method of random effect is considered; otherwise, IVW method of fixed effect will be selected to evaluate the causal effect. Furthermore, we used leave-one-out test to detect outliers and applied funnel and forest plots to examine the pleiotropy intuitively. Finally, a 2-step MR was carried out to estimate the mediated effect of hypertension between PAT and CVDs.^41^ Coefficients analysis was utilized to evaluate indirect effect (β1×β2). β1 and β2 represented the estimated causal effects of PAT on HTN, and HTN on CVDs respectively. Then, Delta method was applied to derive standard error and calculate the proportion of mediated effect in total effect.^42^

To display the estimated causal effect of PAT on CVDs, odds ratio (OR) as well as 95% confidence interval (CI) was employed. All p-values were doubled tailed and corrected by FDR method. Statistical analysis was mainly executed in R project (4.3.1 version) by applying MR-PRESSO and TwoSampleMR packages.

## RESULTS

### Traits of Selected SNPs

After screening P (P < 5 × 10^−8^), eliminating LD (r^2^ < 0.001, 10,000 kb), retrieving in PhenoScanner, and excluding palindromic SNPs, 11 significantly independent SNPs of PAT were extracted as IVs, which summarized 1.32% of the variability (R^2^). F-statistics greater than 10 indicated the strength of the IVs and reflected no weak-instrument bias acquiescently. More details were provided (Table S2 and S3).

### Genetic Effect of PAT on CVDs

The MR results especially for IVW model indicated biologically determined PAT was remarkably correlated with greater risks of CVD (OR 1.15; 95% CI 1.09--1.22, P=1.98*10^-5^), HF (OR 2.62; 95% CI 1.28--5.34, P=0.024), HTN (OR 1.11; 95% CI 1.05--1.17, P=8.53*10^-5^), CHD (OR 3.53; 95% CI 1.55--8.06, P=0.014), IHD (OR 2.31; 95% CI 1.14--4.67, P=0.043), IS (OR 3.18; 95% CI 1.63--6.20, P=0.005), and VTE (OR 1.02; 95% CI 1.00--1.04, P=0.038). However, compared with the controls, no obvious relevance was discovered between genetically determined PAT and other CVDs, including CA (OR 1.44; 95% CI 0.72--2.82, P=0.374), AF (OR 0.99; 95% CI 0.98--1.01, P=0.320), AA (OR 1.15; 95% CI 0.19--6.91, P=0.878), HCM (OR 0.27; 95% CI 5.78*10^-3^--12.61, P=0.577), MI (OR 1.39; 95% CI 0.76--2.55, P=0.374), CES (OR 1.50; 95% CI 0.41--5.39, P=0.577), SVS (OR 3.37; 95% CI 0.97--11.78, P=0.106), LAS (OR 4.05; 95% CI 0.77--21.39, P=0.165), and SAH (OR 0.17; 95% CI 2.67*10^-2^--1.07, P=0.060). Results are presented (Figure 2).

**Figure 2.**
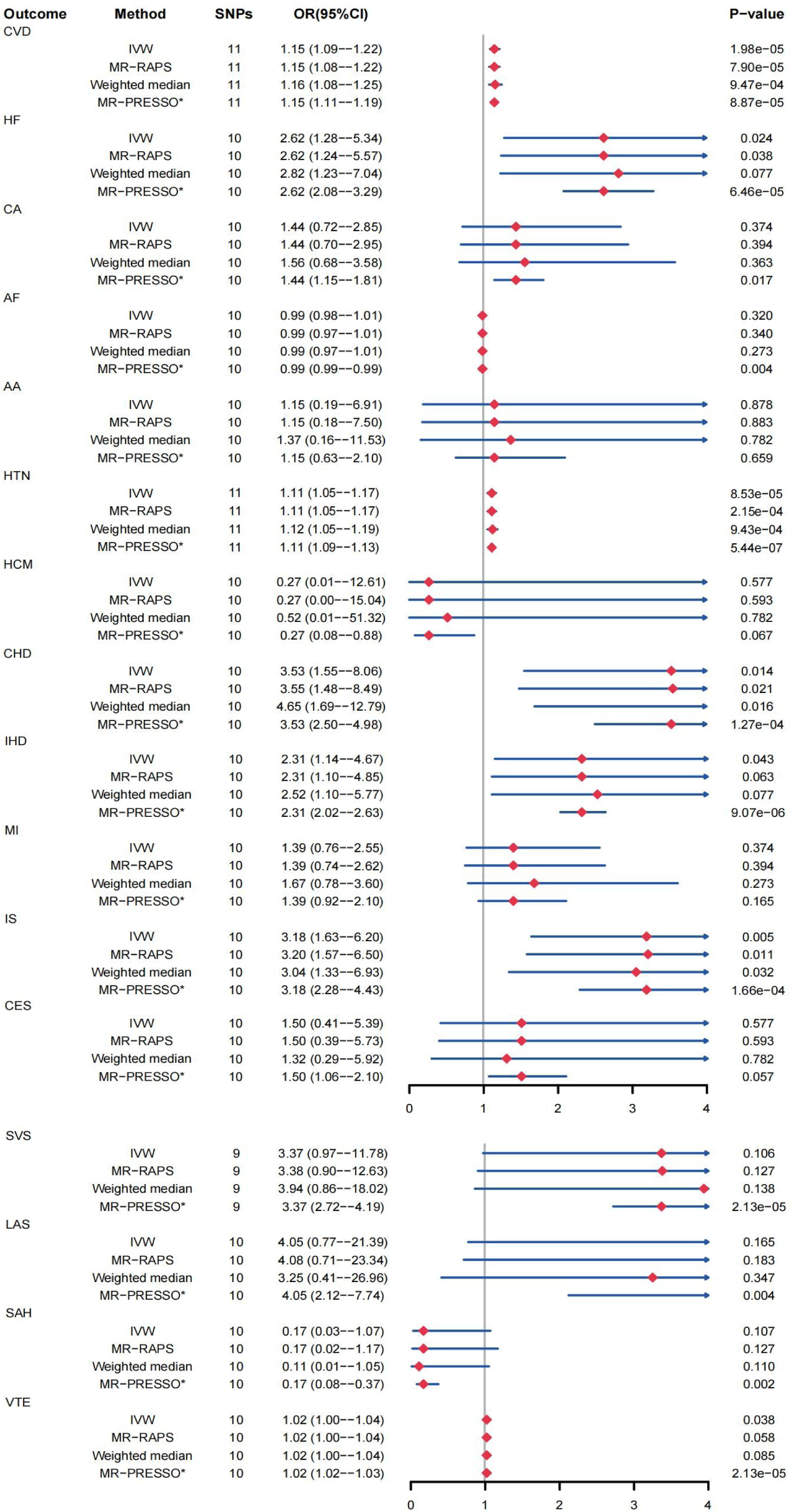
MR estimates of PAT on CVDs. CI, confidence interval; IVW, inverse-variance weighted; MR, Mendelian randomization; MR-RAPS, MR-robust adjusted profile score; MR-PRESSO, MR- pleiotropy residual sum and outlier; OR, odds ratio; SNPs, single nucleotide polymorphisms; *no outlier; AA, aortic aneurysm; AF, atrial fibrillation; CA, cardiac arrhythmias; CES, cardioembolic stroke; CHD, coronary heart disease; CVD, cardiovascular disease; CVDs, cardiovascular diseases; HCM, hypertrophic cardiomyopathy; HF, heart failure; HTN, hypertension; IHD, ischemic heart disease; IS, ischemic stroke; LAS, large artery stroke; MI, myocardial infarction; PAT, pericardial adipose tissue; SAH, subarachnoid hemorrhage; SVS, small vessel stroke; VTE, venous thromboembolism.

### Sensitivity Analysis

The p-value for Q statistics greater than 0.05 instructed no heterogeneity between individual SNP and guided to the fixed-effect IVW model to estimate causal effects chiefly. MR-RAPS and weighted median analyses conducted to detect the causal effect of increased PAT on CVDs were fairly consistent with the IVW model. No significant horizontal multiplicity was discovered based on the intercept of MR-Egger analysis. Besides, no outlying SNPs were identified and excluded using MR-PRESSO analysis. Further, leave-one-out test, along with forest and funnel plots gave an intuitive presence of potential heterogeneity. All results of sensitivity analysis are available (Table S7 and S8).

### 2-step Mediation MR

To figure out what role HTN played between PAT and CVDs, we adopted 2-step MR analysis to assess the intermediary effect. After a bundle of sensitivity analyses, the salient results supported that associations of PAT with CVD, HF, CHD, IHD, and IS were mediated by HTN to some extent. The mediated proportions of HTN were for HF (13.23%; 95% CI 11.47%--15.00%), CHD (13.51%; 95% CI 11.48%--15.55%), IHD (19.18%; 95% CI 17.13%--21.24%), IS (14.51%; 95% CI 12.67%--16.35%). Results are shown (Figure 3 and 4).

**Figure 3.**
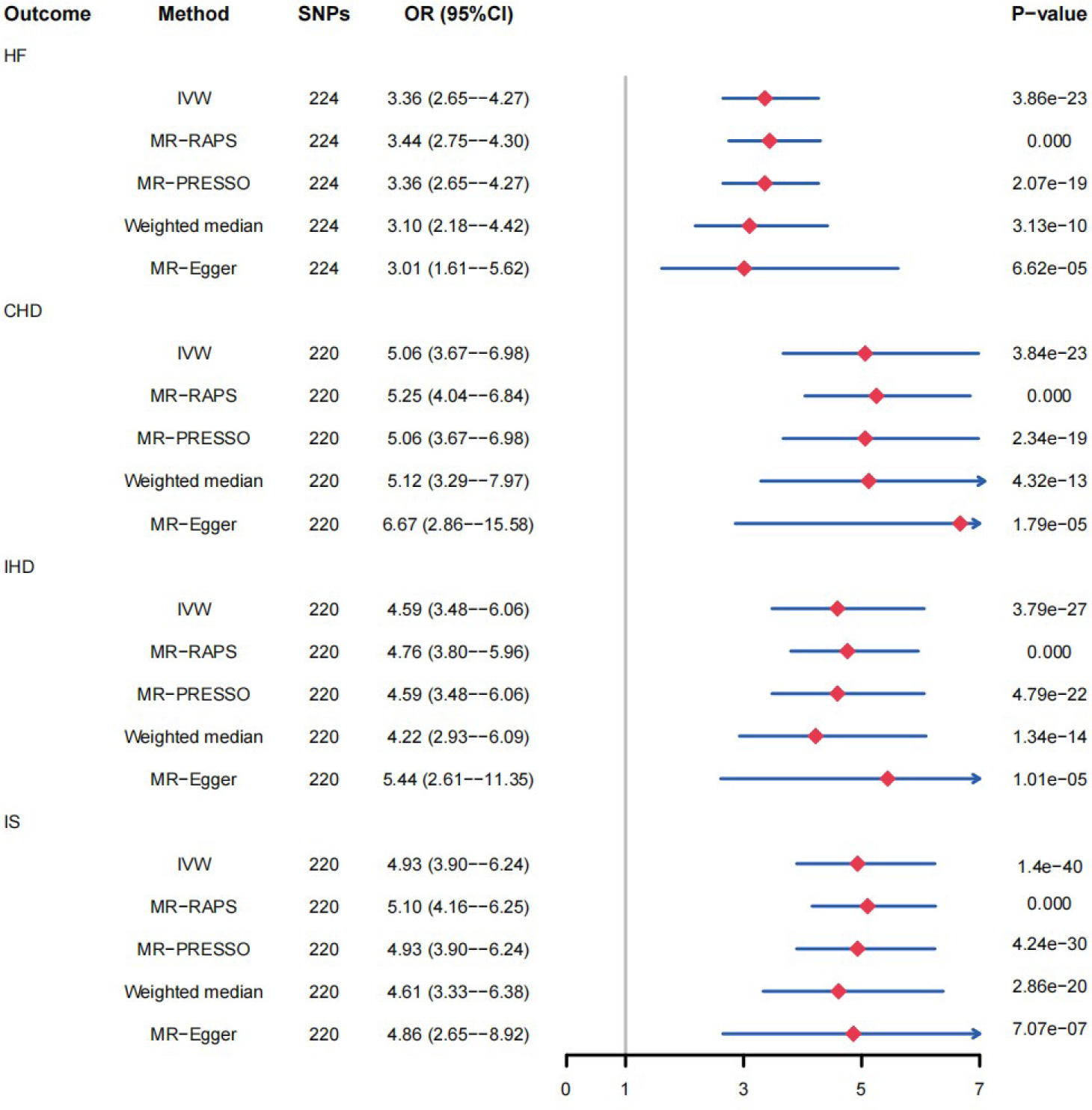
MR estimates of HTN on CVDs. CI, confidence interval; IVW, inverse-variance weighted; MR, Mendelian randomization; MR-RAPS, MR-robust adjusted profile score; MR-PRESSO, MR- pleiotropy residual sum and outlier; OR, odds ratio; SNPs, single nucleotide polymorphisms; CHD, coronary heart disease; CVDs, cardiovascular diseases; HF, heart failure; HTN, hypertension; IHD, ischemic heart disease; IS, ischemic stroke.

**Figure 4.**
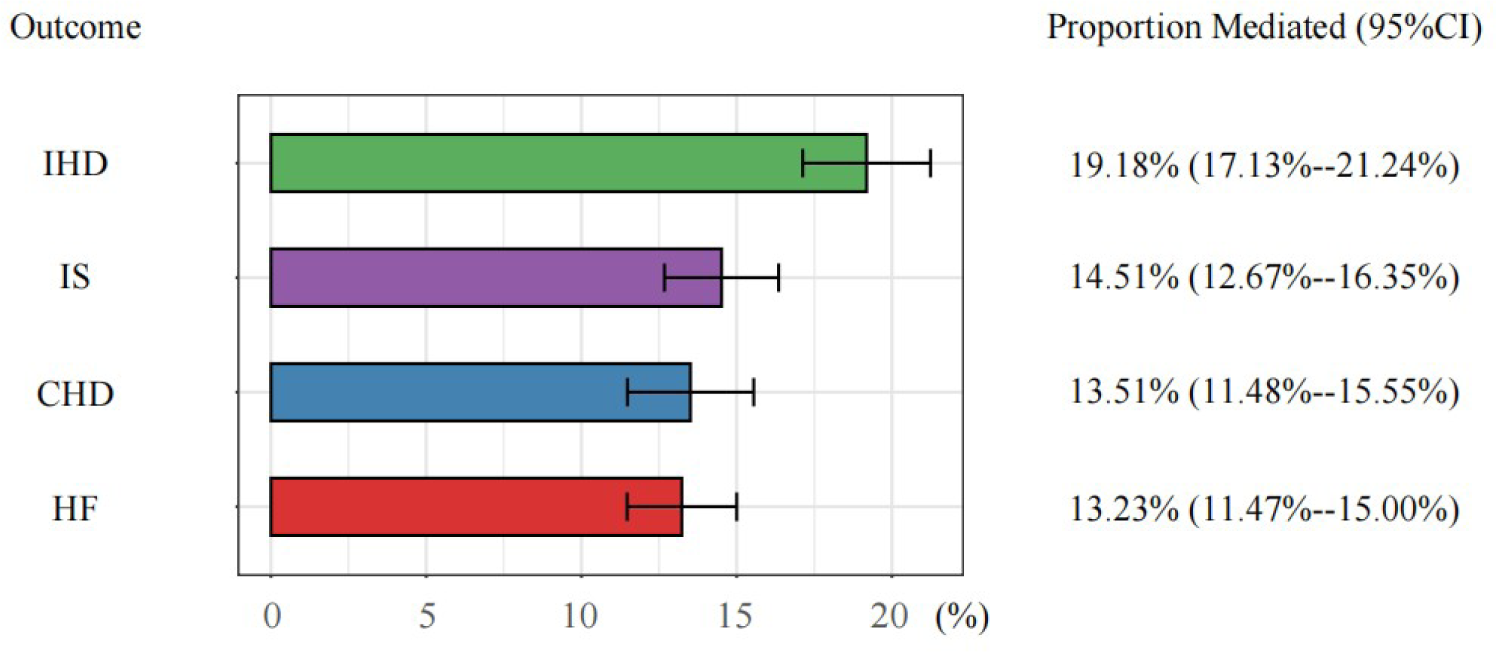
MR estimates of proportions mediated by HTN between PAT and CVDs. CI, confidence interval; MR, Mendelian randomization; CHD, coronary heart disease; CVDs, cardiovascular diseases; HF, heart failure; HTN, hypertension; IHD, ischemic heart disease; IS, ischemic stroke; PAT, pericardial adipose tissue.

## DISCUSSION

In the MR study, IVs containing 11 SNPs were designated to assess the causal effect. The result suggested that biologically determined PAT was significantly correlated with greater risks of CVD, HF, HTN, CHD, IHD, IS, and VTE after standard MR and sensitivity analyses. Hypertension was proved to play a strong mediating role in the linkage. But limited evidence supported a potential causal association between PAT and CA, AF, AA, HCM, MI, CES, SVS, LAS, SAH risks.

Classical anthropometric metrics including body mass index have limitations in evaluating the correlation between PAT and CVDs.^43,44^ Nowadays, amounts of studies have addressed the association. Satish et al. revealed that elevated PAT volume came with a greater risk of HF with preserved ejection fraction after adjusting for abdominal visceral adipose tissue.^16^ Pugliese et al. demonstrated patients suffering from HF of reduced ejection fraction were observed with significantly decreased PAT thickness compared to controls.^45^ But the lack of GWAS data prevented us estimating the effect of PAT on HF with different subtypes. A meta-analysis declared patients with AF were significantly associated with increased PAT volume.^46^ Iacobellis G et al. highlighted PAT thickness quantified by echocardiography was related to the seriousness of CAD.^47^ According to Bryan et al., the hazard of MI conferred with increased PAT thickness measured by computed tomography.^48^ However, some reports presented contradictory results. Park J et al. demonstrated no remarkable relationship was observed between PAT and AF.^49^ An observational study presented PAT was uncorrelated with CAD.^50^ Also, research confirmed that an upper component in PAT promoted the recovery of cardiac function after MI.^51^ Although traditional transect studies are vulnerable to inverse causation bias and confounders, these inconsistent observational outcomes still confuse us. Looking forward to more high-quality studies to give an affirmative answer. The association between PAT and CVDs worth exploring further.

In general, the MR research revealed that PAT was an expected predictor for increased risk of CVDs. PAT, as a specific type of visceral fat, is related to metabolic syndrome and hazard of CVDs.^52^ And amounts of inflammatory and bioactive factors, including adipocytokines, Interleukin-6, and tumor necrosis factor-α could be secreted by PAT through paracrine and vasocrine functions, which promoted cardiovascular disease progression.^53,54^ Unexpectedly, a strong connection was discovered between PAT and HTN. Known as a major hazard for CVDs,^7^ it was reasonable to conjecture that HTN played an important intermediary role between PAT and CVDs. The mediating effect was confirmed by 2-step mediation MR analysis, which might point the way to understanding how PAT facilitated CVDs occurrence. Recent studies have declared glucagon-like peptide-1 analogue and sodium-dependent glucose transporters 2 inhibitors could diminish PAT accumulation.^55,56^ These above paint such a picture of prospect that PAT might act as a potential drug target to provide cardiovascular benefits and antihypertensive drugs could be conjunctly used in CVDs progression. Nowadays, obesity and cardiovascular diseases are heavy burdens on global health. Specific and effective drugs targeting PAT would offer significant health consequences, especially for people with obesity who were more susceptible to developing CVDs because of greater PAT accumulation.

But as far as the causal effect of PAT on AF, our results are discrepant with most previous research. Several possible explanations can be made for this inconsistency. Firstly, whether PAT is an independent impact factor for atrial fibrillation is still critical.^46,49^ Secondly, the occurrence of AF is genetically and environmentally driven, but we assessed the association between PAT and AF only genetically. Besides, MR research usually considers long-term impacts. Thirdly, confounding factors are unignorable in previous observational studies, such as hyperlipidemia, and drug utilization. And the poor reproducibility of PAT thickness measured by echocardiography may account for part of these differing discoveries.^57^ In addition, cross-sectional and observational studies have their limitations and shortcomings. Here, we expect for more prospectively longitudinal and experimental research with greater persuasive power. Finally, no matter whether as a contributor or follower or a double-edged sword of both protection and destruction, the role played by PAT in CVDs remains to be further confirmed in humans.

Our research has several advantages. First of all, it is the first MR research to detect the prospective causal association between genetically determined PAT and multiple CVDs. Next, the latest and large-scale GWAS datasets used in the MR study provide a more persuasive way to assess causal relationship. Furthermore, diverse sensitivity analyses, MR methods, and IVs strength evaluation were conducted to validate the credibility of outcomes and avoid weak-tool bias as well as traditional confounding factors.

Of note, there are some limitations in the MR research. Firstly, due to the lack of original GWAS data of PAT, we could not further carry out multi-variable MR to evaluate the causal effects deeply and control potential confounders, such as BMI, visceral adipose tissue, triglyceride, and hypertension. Secondly, participants in the MR study are all European, which means these findings do not necessarily apply to another population. Thirdly, some degree of sample overlap appeared between PAT and CVDs, the maximum possible percentages of which were for CVD (5.81%), AF (6.08%), HTN (5.81%), MI (6.10%), SAH (5.95%), and VTE (7.80). But Pierce has revealed that 10% of IVs still retain 90% of analytical power.^58^ And the corresponding F-statistics were large enough to compensate for its effect to some extent. Fourthly, in view of insufficient statistical efficacy, susceptibility to outlying SNPs, and larger standard deviation than other methods, the MR-Egger sometimes presented inconsistent results. Otherwise, although a series of sensitivity analyses were adopted, potential multiplicity and bias might still remain. Finally, the cases of AA (2825), HCM (556), LAS (4373), SAH (1693), and VTE (4620) were fairly small when compared with other outcomes.

## CONCLUSION

MR research indicated biologically determined PAT was significantly correlated with greater risks of CVD, HF, HTN, CHD, IHD, IS, and VTE. The study demonstrates the potential that PAT serves as a drug target for several cardiovascular diseases. But the correlation between PAT and multiple CVDs deserves further exploration.

## PERSPECTIVES

Our study discovered the causal effect of PAT on CVDs and HTN played a vital intermediary role in it. PAT demonstrates significant potential as a therapeutic target for CVDs, and antihypertensive strategies might be taken into account. Usually, people with obesity get additional PAT cumulation and are more prone to CVDs. In terms of obesity and CVDs being heavy burdens on global health, drugs specifically targeting PAT will offer huge health benefits. However, the pathophysiologic mechanism between PAT and CDVs remains unclear, more scientifically-designed studies are required to clarify the relationship.

## Data Availability

All data of this research are available.

## Nonstandard Abbreviations and Acronyms

AA: aortic aneurysm
AF: atrial fibrillation
CA: cardiac arrhythmias
CAD: coronary artery disease
CES: cardioembolic stroke
CHD: coronary heart disease
CI: confidence interval
CVD: cardiovascular disease
CVDs: cardiovascular diseases
GWAS: genome-wide association study
HCM: hypertrophic cardiomyopathy
HF: heart failure
IHD: ischemic heart disease
IS: ischemic stroke
IVs: instrumental variables
IVW: inverse-variance weighting
LAS: large artery stroke
LD: linkage disequilibrium
MI: myocardial infarction
MR: mendelian randomization
OR: odds ratio
PAT: pericardial adipose tissue
SAH: subarachnoid hemorrhage
SNPs: single-nucleotide polymorphism
SVS: small vessel stroke
VTE: venous thromboembolism.

## Acknowledgments

Feng Xie and Shai-hong Zhu made a contribution to the conception. Feng Xie and Peng Cao drafted this paper. Hai-bo Tang, Zhi Luo and Li-yong Zhu gave a critical revision. Li-yong Zhu obtained funding. Sincere thanks to all contributors of utilized GWASs.

